# Antibody titers before and after booster doses of SARS-CoV-2 mRNA vaccines in healthy adults

**DOI:** 10.1101/2021.11.19.21266555

**Authors:** Alexis R. Demonbreun, Amelia Sancilio, Lauren A. Vaught, Nina L. Reiser, Lorenzo Pesce, Elizabeth M. McNally, Thomas W. McDade

**Author notes:** **Corresponding authors** Thomas McDade, PhD, Northwestern University, 1810 Hinman Avenue, Evanston, IL 60208, Alexis Demonbreun PhD, Northwestern University, 303 Superior SQ 5-512, Chicago, IL 60611.

## Abstract

Two-dose messenger RNA vaccines (BNT162b2/Pfizer and mRNA-1273/Moderna) against SARS-CoV-2 were rolled out in the US in December 2020, and provide protection against hospitalization and death from COVID-19 for at least six months. Breakthrough infections have increased with waning immunity and the spread of the B.1.617.2 (Delta) variant in summer 2021, prompting approval of boosters for all adults over 18. We measured anti-receptor binding domain (RBD) IgG and surrogate virus neutralization of the interaction between SARS-CoV-2 spike protein and the human angiotensin-converting enzyme (ACE2) receptor, before and after boosters in N=33 healthy adults. We document large antibody responses 6-10 days after booster, with antibody levels that exceed levels documented after natural infection with COVID-19, after two doses of vaccine, or after both natural infection and vaccination. Surrogate neutralization of B.1.617.2 is high but reduced in comparison with wild-type SARS-CoV-2. These data support the use of boosters to prevent breakthrough infections and suggest that antibody-mediated immunity may last longer than after the second vaccine dose.

## Introduction

Two-dose messenger RNA vaccines (BNT162b2/Pfizer and mRNA-1273/Moderna) against SARS-CoV-2 provide protection against hospitalization and death from COVID-19 for at least six months^1,2^. Breakthrough infections have increased with waning immune protection and the spread of the B.1.617.2 (Delta) variant in summer 2021, prompting US regulatory approval of boosters for higher risk individuals. Serological studies reveal modest antibody responses to the first priming dose of mRNA vaccine with substantially stronger responses to the second dose. The magnitude of antibody response to booster doses, more than 6 months after full vaccination, is not known. Understanding antibody responses to COVID-19 boosters is important for forecasting the level and duration of antibody-mediated protection.

## Methods

All research activities were implemented under protocols approved by Northwestern University’s institutional review board (#STU00212457 and #STU00212472).

Participants e-consented and completed online surveys regarding COVID-19 viral history and vaccination status. Finger stick dried blood spot (DBS) samples were self-collected prior to booster administration and 6-10 days after receiving mRNA booster vaccine (paired samples). Results were compared with data from a prior community-based study using the same protocols, which quantified antibody responses after diagnosis of COVID-19 or administration of dose 2 of mRNA COVID-19 vaccine^3^. Participants were classified as seronegative or seropositive based on the presence of anti-receptor binding domain (RBD) IgG prior to vaccination.

Anti-RBD IgG ELISA protocol was performed as described, indexing anti-RBD IgG to a CR3022 calibration curve^3,4^. Surrogate virus neutralization (SVN) of spike-human angiotensin-converting enzyme (ACE2) receptor was performed as described ^5^, with an initial 1:40 dilution and half maximal inhibitory concentration (IC50) from 1:2 serial dilution.

Statistics were performed using Wilcoxon, Kruskal-Wallis, or Spearman r where appropriate. p<0.05 was considered statistically significant.

## Results

Participants (N=33, female=16) were fully vaccinated 182-290 days (mean = 237.9) prior to booster. Median pre-booster anti-RBD IgG was 4.4µg/mL, with no significant differences between females and males (4.3 vs. 4.4µg/mL; z=0.414; p=0.68) and pre-booster IgG negatively associated with age (Spearman r=-0.69, p<0.0001) (median age 43 yrs). IgG increased ∼25-fold after booster, with median concentration of 101.6µg/mL (**Figure 1A and 1B**). Females had non-significantly higher post-booster IgG than males (111.3 vs. 79.5 µg/mL; z=1.33; p=0.18), and post-booster IgG was negatively associated with age (Spearman r=-0.44, p=0.01).

**Figure 1.**
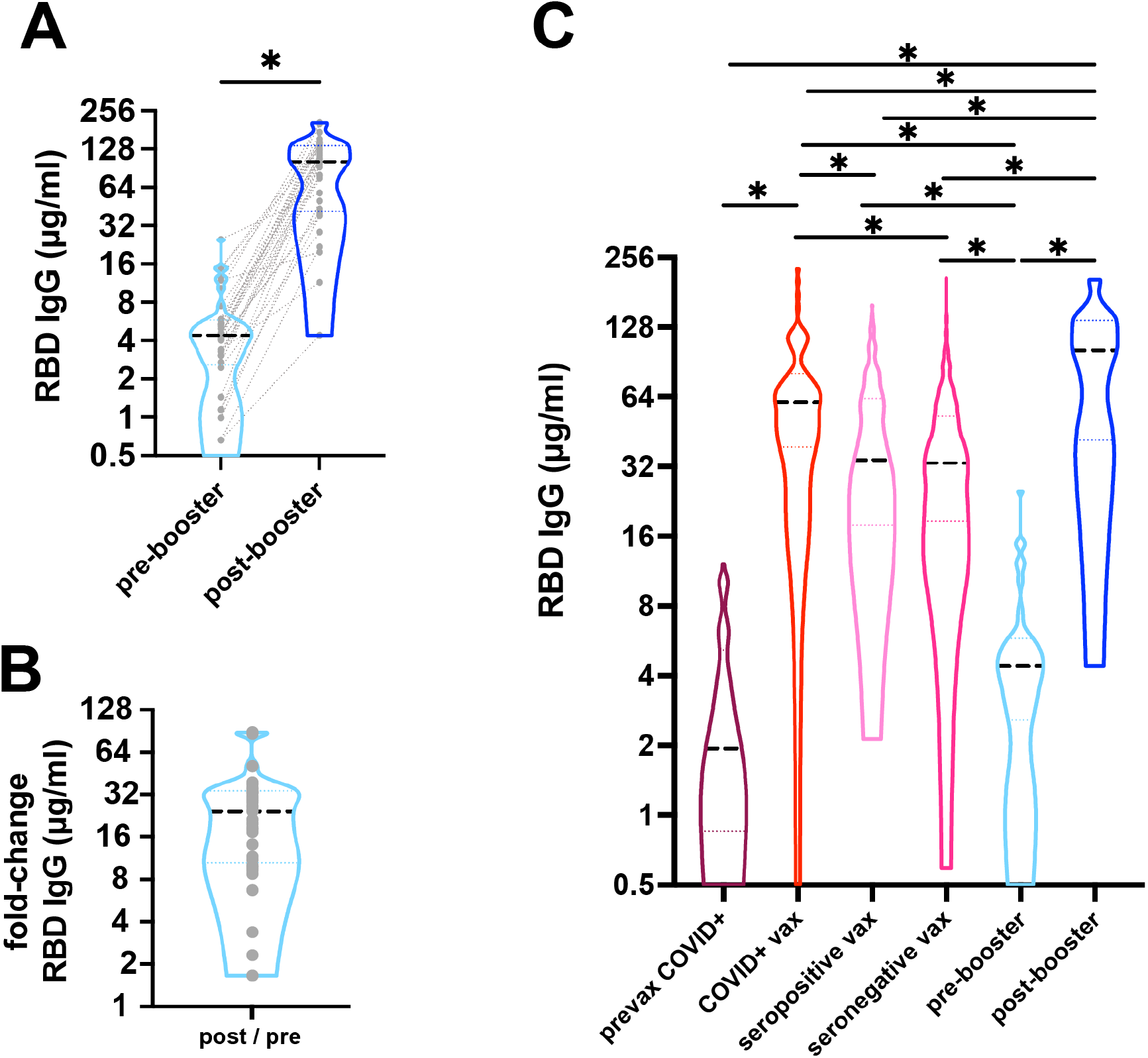
Robust IgG antibody response after COVID-19 mRNA booster vaccination. **A)** Response to COVID-19 mRNA vaccine and booster was measured as anti-RBD IgG antibodies from dried blood spots. Median IgG concentration (black dashed line) increased from 4.4µg/ml pre-booster to 101.6µg/ml post-booster (*p<0.001). Grey dotted lines represents paired samples. n=33. **B)** There was a median 25-fold change post-booster. **C)** Median anti-RBD IgG concentration (black dashed line) are shown. Individuals with outpatient COVID-19 had a median of 1.92 µg/ml (n=76) 14-42 days after infection, while individuals with a history of COVID-19 followed by vaccination were higher (60.61µg/ml, n=73, 5-42 days after 2^nd^ dose). Individuals without a known history of COVID-19 who were either seropositive or seronegative and then 2-dose vaccinated had median IgG of 34.15µg/ml (n=181) and 33.09µg/ml (n=687), respectively. Pre-booster levels mean 237.9 days after 2-doses of vaccine were 4.4 µg/ml (n=33) compared to post-booster vaccination level of 101.6 µg/ml (n=33). Dotted lines represent the 25^th^ and 75^th^ percentiles. (*p<0.001).

Median IgG concentration after booster was significantly higher than levels after clinical diagnosis of COVID-19+ (1.92 µg/ml), or after two doses of mRNA vaccine with and without a history of prior COVID-19 (COVID+ 60.61, seropositive 34.15, seronegative 33.09 µg/ml; p<0.0001) (**Figure 1C**). All participants responded to booster with an increase in antibody concentration, but the magnitude of response (post-dose – pre-dose IgG) ranged from 3.74 to 195.6 µg/mL. Pre-dose antibody levels were strong predictors of post-dose levels (Spearman r=0.53, p=0.0015).

Surrogate virus neutralization responses were similar. The pre-booster SVN of wildtype spike-ACE2 interaction was low, with only four participants having detectable neutralization at IC50 titer > 40. Post-booster median IC50 was 283.9, with values as high as 647.5 (**Figure 2A**). SVN IC50 did not differ by sex (310.6 vs. 273.6, z=0.18, p=0.86). However, post-IC50 negatively associated with age (Spearman r=-0.35, p=0.045). SVN post-booster was highly correlated with anti-RBD IgG post-booster (Spearman r=0.83, p<0.0001) (**Figure 2B**).

**Figure 2.**
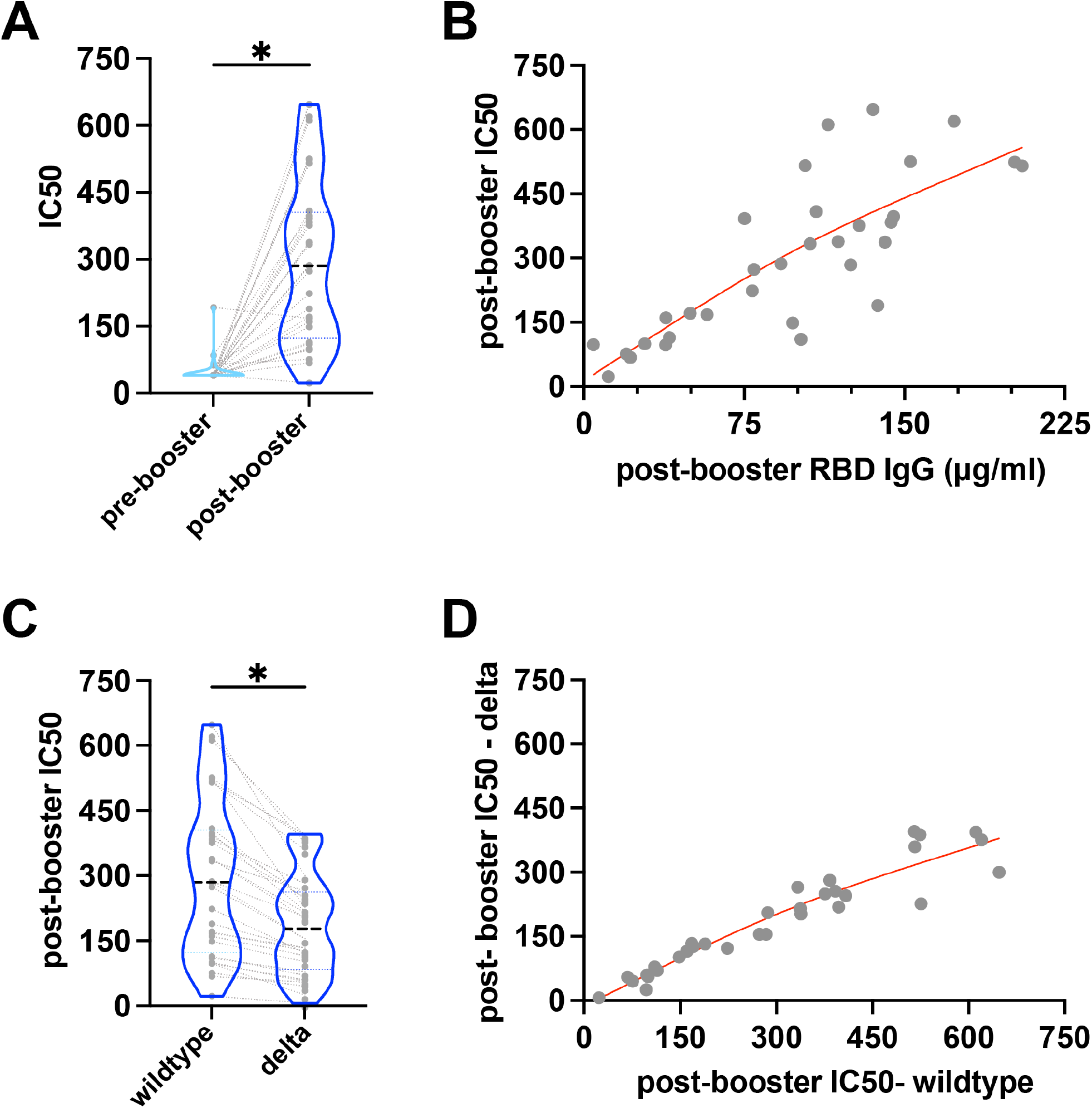
Robust neutralizing antibody response after COVID-19 mRNA booster vaccination. Dried blood spots were evaluated after COVID-19 mRNA booster vaccine for surrogate neutralizing capacity. **A)** Median IC50 titers of surrogate virus neutralization of wildtype spike-ACE2 interaction was significantly higher post-booster (283.9) than pre-booster (40) (*p<0.0001). **B)** Post-booster anti-RBD levels positively correlated with post-booster IC50 titers (Spearman r=0.83, p<0.0001, n = 33). Redline denotes spline. **C)** Median post-booster IC50 titers were significantly lower against delta B.1.617.2 (154.9) compared to wildtype titers (283.9) (*p<0.0001, n = 33). **D)** Post-booster wildtype IC50 titers positively correlated with post-booster delta IC50 titers (Spearman r=0.95, p<0.001, n = 33). Redline denotes spline.

Post-booster SVN of B.1.617.2 variant was significantly lower than wildtype SARS-CoV-2 (median IC50 154.9 vs. 283.9, z=4.99, p<0.000) (**Figure 2C**). There were no differences between females and males in B.1.617.2 neutralization (178.5 vs. 154.8, z=0.13, p=0.90), while IC50 negatively associated with age (Spearman r=-0.45, p=0.008). Post-dose IC50 wildtype titer was a strong predictor of post-dose IC50 B.1.617.2 titer (Spearman r=0.95, p<0.001) (**Figure 2D**).

## Discussion

Results from this study indicate that BNT162b2/mRNA-1273 boosters generate large antibody responses in healthy adults, with post-booster antibody levels that exceed levels documented after natural infection with COVID-19, after two doses of vaccine, or after both natural infection and vaccination. Surrogate neutralization of B.1.617.2 is high but reduced in comparison with wild-type SARS-CoV-2. These data support the use of boosters to prevent breakthrough infections^6^ and suggest that antibody-mediated immunity may last longer than after the second vaccine dose.

Study limitations include small sample size, short timeframe, and absence of cellular immunity measures. While waning antibodies may contribute to breakthrough infections, it is important to investigate the effects of boosters on cell-mediated immunity.

## Data Availability

Requests for the potentially identifiable dataset should be made by qualified researchers trained in human subject confidentiality protocols to Dr. Thomas McDade at Northwestern University.

## Acknowledgements

Supported by the National Science Foundation 2035114, NIH 3UL1TR001422-06S4, Northwestern University Office of Research, and a generous gift from Dr. Andrew Senyei and Noni Senyei. The funding sources had no role in the study design, data collection, analysis, interpretation, or writing of the report.

